# How well can we forecast the COVID-19 pandemic with curve fitting and recurrent neural networks?

**DOI:** 10.1101/2020.05.14.20102541

**Authors:** Zhuowen Zhao, Kieran Nehil-Puleo, Yangzhi Zhao

## Abstract

Predictions of the COVID-19 pandemic in USA are compared using curve fitting and various recurrent neural networks (RNNs) including the standard long short-term memory (LSTM) RNN and 10 types of slim LSTM RNNs. The curve fitting method predicts the pandemic would end in early summer but the exact date and scale vary with the evolving data used for fitting. All LSTM RNNs result in short-term (8 to 10 days) predictions with comparable accuracies (smaller than 10 %) to curve fitting—they do not show advantage over curve fitting.

## 1. Introduction

Since the first report in late January, COVID-19 has spread to the whole world and become a global pandemic. The accurate prediction of the scale and time of the pandemic is helpful information to the the public to cope with the crisis. In this paper, we compare the predictions with two methods, i.e., curve fitting of a modified exponential function and recurrent neural network (RNN). The curve fitting method is basically an extrapolation based on a well-fitted analytical function that can be used for full-range predictions (including short-term predictions up to any time before end). If no noise added, the function or its derivative (delta values if not differentiable) is not expected to capture local fluctuations. RNN is typically used for predicting serial events, which is potentially capable for capturing daily cases fluctuations of the COVID-19 pandemic. This paper aims at (i) evaluating full-range predictions with curve fitting as well as at (ii) comparing short-term (8–10 days) forecasting with smooth curve fitting (no noise added) and RNN.

In order to overcome the possible long-term learning difficulty due to gradient vanishing or gradient explosion of simple RNN, long short-term memory (LSTM) technique is usually adopted [1]. In addition to the standard LSTM RNN, we also experiment with simplified (slim LSTM) networks—less parameters in the gate (8 types) and memory cell (2 types) structures [2, 3].

## 2. COVID-19 data

Data source is located at Github repository https://github.com/CSSEGISandData/COVID-19github.com/CSSEGISandData/COVID-19, which is originally from Johns Hopkins University Coronavirus Resource Center. It has confirmed patients and deaths data in time series in the category of regions and the corresponding countries. We plot confirmed patients from Hubei province of China (first report location), China, USA, and Italy with respect to the days since first case reported in each region shown in Fig. 1. Italy curve is shorter than the other regions because the first patient reported for Italy was on January 31 while it was January 22 for both China and USA.

**Figure 1:**
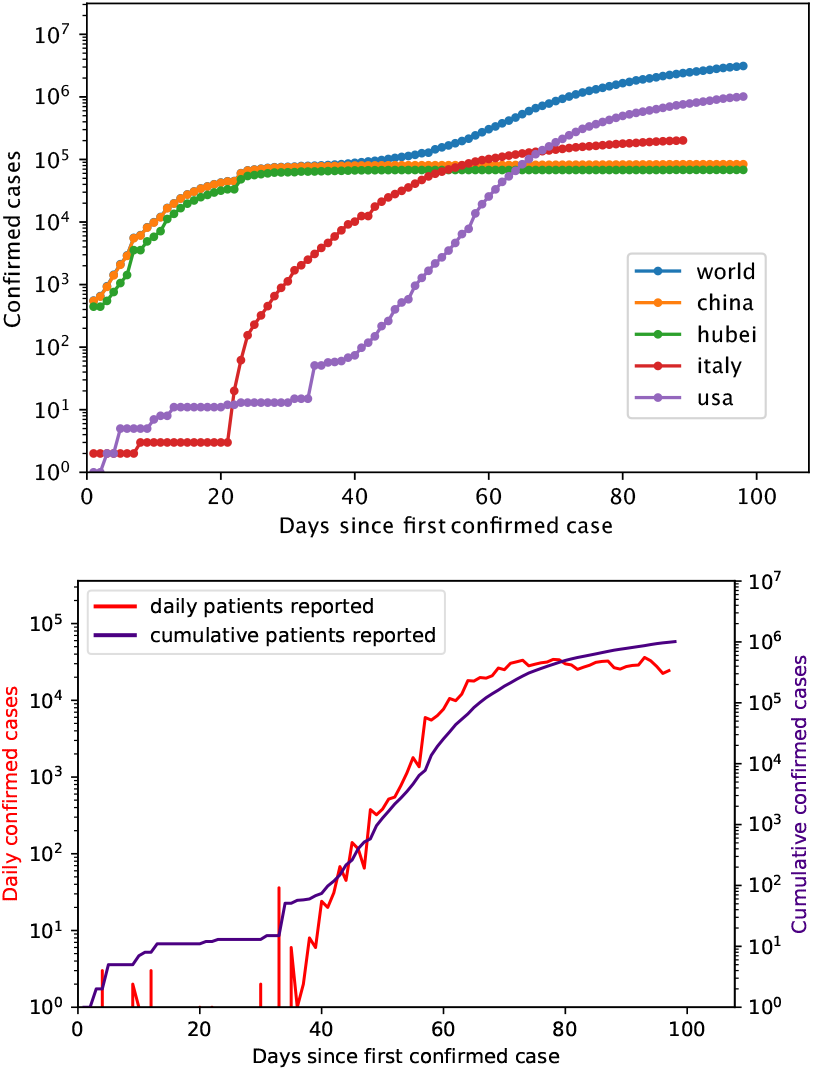
Top: confirmed COVID-19 cases of the whole world and four regions since first case was reported. Bottom: cumulative patients and new cases reported per day of USA since the first case was reported. Data in both plots are up to April 28.

## 3. Full-range prediction with curve fitting

Langel [1] reported a model in the form of a modified exponential function Eq. (1) that describes the evolution of the cumulative cases of the pandemic.

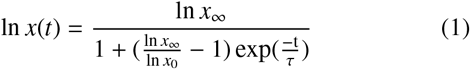

,where *x*_∞_, *x*_0_, τ are final fitting parameter (ultimate number of cases), initial fitting parameter (for numerical purpose), and a parameter that controls the “curvature” of the curve. The idea behind this model is that the cumulative patients/death curve in principle follows the trajectory of exponential function whose derivative (daily increment) looks like a skewed normal distribution curve.

The model is fitted (least square) with cumulative patients data of three different lengths (82, 88, 98-day) to investigate its performance with evolving data. Table 1 shows the three optimized parameter sets and one parameter set for death data.

**Table 1:** Optimized parameters *x*_∞_, *x*_0_, τ based on cumulative patients (82, 88, 98-day) data and death (60-day) data in USA. parameters curve fitting (normalized RMSE 0.02)

| parameters | cumulative patients |  |  | death |
| --- | --- | --- | --- | --- |
|  | 82-day | 88-day | 98-day | 60-day |
| $x_0$ | 1.1 | 1.2 | 1.6 | 6.8 |
| $x_{\infty}$ | $9.7 \times 10^5$ | $1.0 \times 10^6$ | $1.3 \times 10^6$ | $7.7 \times 10^4$ |
| $\tau$ | 1.0 | 1.1 | 1.3 | 1.2 |

The metric to evaluate the goodness of the fitting, i.e., *normalized* root mean square error (RMSE) is calculated based on true values and the fitted values within the same time frame— their RMSE divided by the mean of true values. The *normalized* RMSE is only 0.04 (or 4 %) by fitting with confirmed infection data and 0.02 by fitting with death data up to April 28 (98-day). The daily patients curve (red dashed line in Fig. 2 top)—delta values of the fitted function—suggests the pandemic in USA would end around June 30, 2020. The daily death curve (blue dashed line in Fig. 2 bottom) suggests the America would see zero death since July. Unfortunately, the model also predicts about 1.3 million infections and 77 thousand deaths in America at the end of COVID-19 pandemic based on the 98-day data. However, the ultimate number of patients by prediction increases from 1.0 to 1.3 million with more data (16 days), see Table 1. This suggests the COVID-19 is a quickly evolving situation, thus one-time full-range prediction may not be very reliable using the curve fitting.

**Figure 2:**
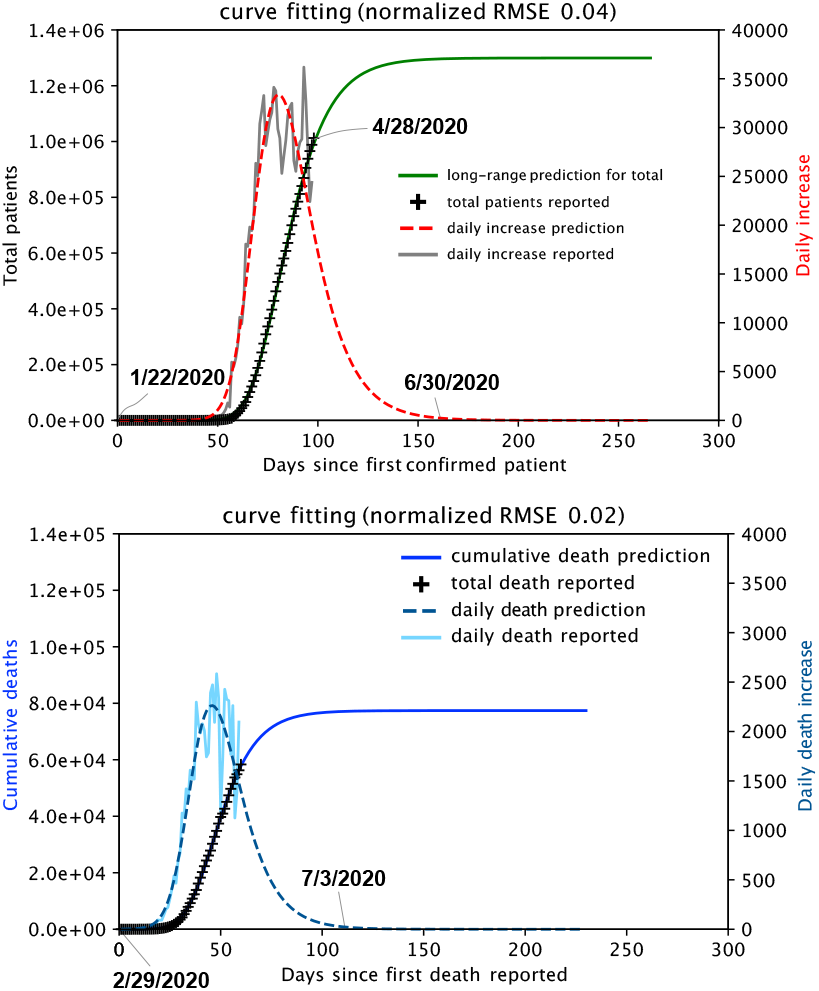
Prediction of total patients (top) and deaths (bottom) due to COVID-19 in USA using data up to April 28 (98 days for patient data and 60 days for death data). The first reported death in USA was on February 29 according to the JHU dataset (this date varies with other media).

## 4. Short-term prediction with curve fitting and LSTM RNNs

### 4.1. Short-term prediction with curve fitting

The dataset is split into training/fitting (90 %) and testing(10 %) subsets (to evaluate the short-term prediction). The function is firstly fitted with the training set to get optimized parameters. Then the prediction from extrapolation are evaluated using *normalized* RMSE against testing set. Figure 3 shows the short-term predictions (9-day and 10-day) using 82-day and 88-day data for fitting respectively. The *normalized* RMSE values vary with different fitting data lengths and they are both smaller than 10 %. Moreover, curve fitting tends to underestimate cases for short-term prediction.

**Figure 3:**
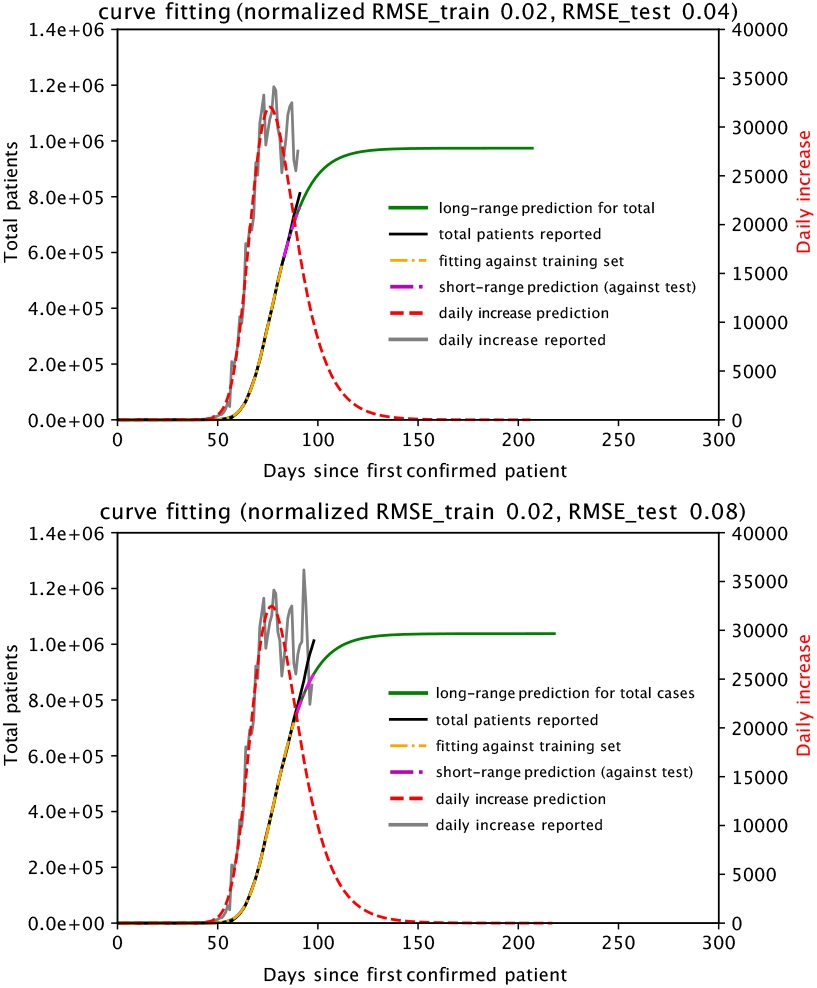
Short-term prediction of COVID-19 patients in USA by fitting with 82-day (top) and 88-day data (bottom).

### 4.2. Short-term prediction with standard LSTM RNN

The standard LSTM RNN is built on Keras with TensorFlow as the backend software. The network has an architecture of four LSTM cells followed by a dense output layer. Figure 4 shows the structure for one of the LSTM cells. Hyperbolic tangent (tanh) function (default in Keras library) is used as activation function for all gates and memory cells. We also tested sigmoid function, another popular choice for activation function in RNN, but the prediction was totally off with the same network architecture. “Adam optimizer” from Keras library is adopted for the backpropagation updates and mean squared error is used as the loss function. The formula of standard LSTM RNN are summarized in the following[4, 5].

**Figure 4:**
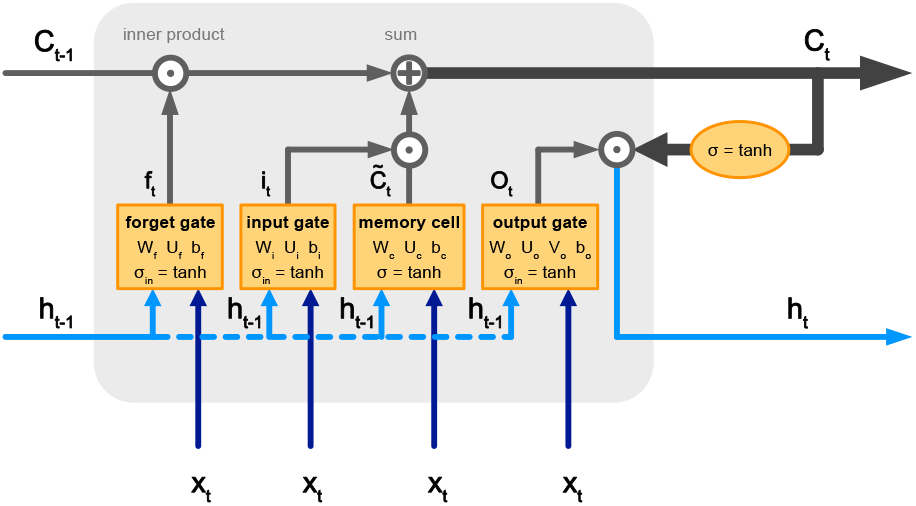
Schematics of LSTM RNN structure.

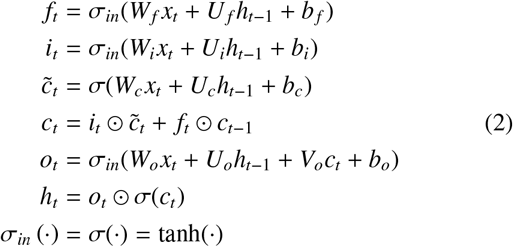

, where *f_t_, i_t_, o_t_, c_t_* denotes forget, input, output gate, and cell structure at time t in LSTM cells respectively. *h_t_* is state variable at time t. *W_f_*, *W_i_, W_o_, W_c_, U_f_*, *U_i_, U_o_, U_c_, V_o_* are weight matrices and *b_f_*, *b_i_, b_o_, b_c_* are bias vectors. σ_*in*_(·) (for gates, see Fig. 4) and σ(·) are activation functions.

The short-term predictions are made using dataset of two different lengths (91 and 98 days) respectively. Each dataset is normalized into the range of 0 to 1. Four new subsets *X*_train_, *Y*_train_, *X*_test_, and *Y*_test_ are made from each dataset, among which *X*_train_ and *Y*_train_ are used to train the RNN network, and *Y*_test_ is used for evaluating the predictions from *Y*_train_. Take 91-day data for example, data points in the first 82 days are used as training set (90 %) and the rest (9 days) are used as testing set (10 %). From the training set, *X*_train_ and *Y*_train_ are made by removing the last day and the first day from the original training set respectively, such that *Y*_train_ is one day ahead of *X*_train_ for each data point. In other words, *Y*_train_ contains true values of the one-day evolution of *X*_train_ (both subsets have 81 days). *X*_test_ and *Y*_test_ are made in the same way from the training test (8 days).

Figure 5 shows evaluation results against the training and testing sets. The *normalized RMS E train* and *RMS E test* demonstrate how well the RNN network is trained and how well the trained network forecasts respectively. It is seen from Fig. 5, both evaluations are of the same level of magnitude compared to curve fitting.

**Figure 5:**
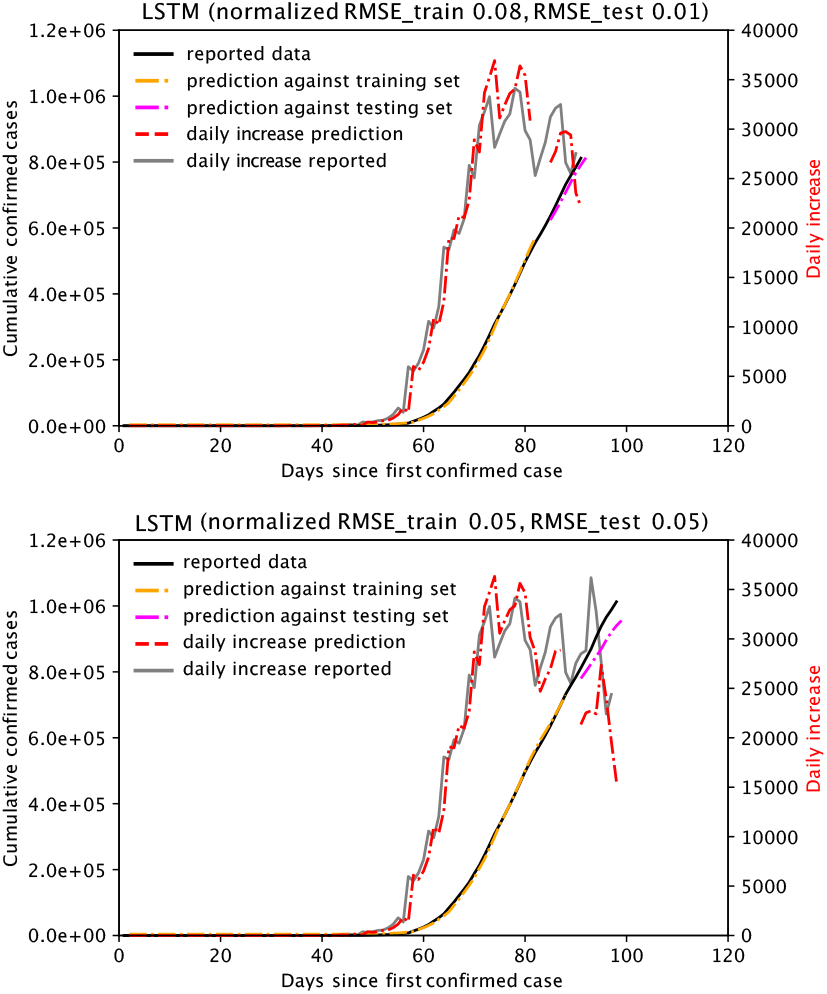
Prediction of cumulative patients in USA with LSTM using 82-days (top) and 88-day data (bottom).

### 4.3. Short-term prediction with slim LSTM RNN

LSTM RNN can be simplified by reducing parameters in the three gates (forget, input, and output gate) or in memory cells[2, 3], which is expected to accelerate the convergence. There are 10 types of simplified LSTM RNN used in this study, which are denoted as LSTM1, LSTM2, LSTM3, LSTM4, LSTM4a, LSTM5, LSTM5a, LSTM6, LSTM10, LSTM11 respectively. The detailed formula for each type can be found in Section 7 Appendix.

Figure 6 shows the prediction of patients in USA with slim LSTM RNN(s) using 82-day data. They all have same level of accuracies compared to standard LSTM and curve fitting, among which LSTM6 demonstrates the best prediction accuracy (least *normalized* RMSE against testing set). All slim LSTM RNN networks tend to underestimate the future cases compared to the reported data (magenta dash-dotted line is below the black solid line for all models in Fig. 6).

**Figure 6:**
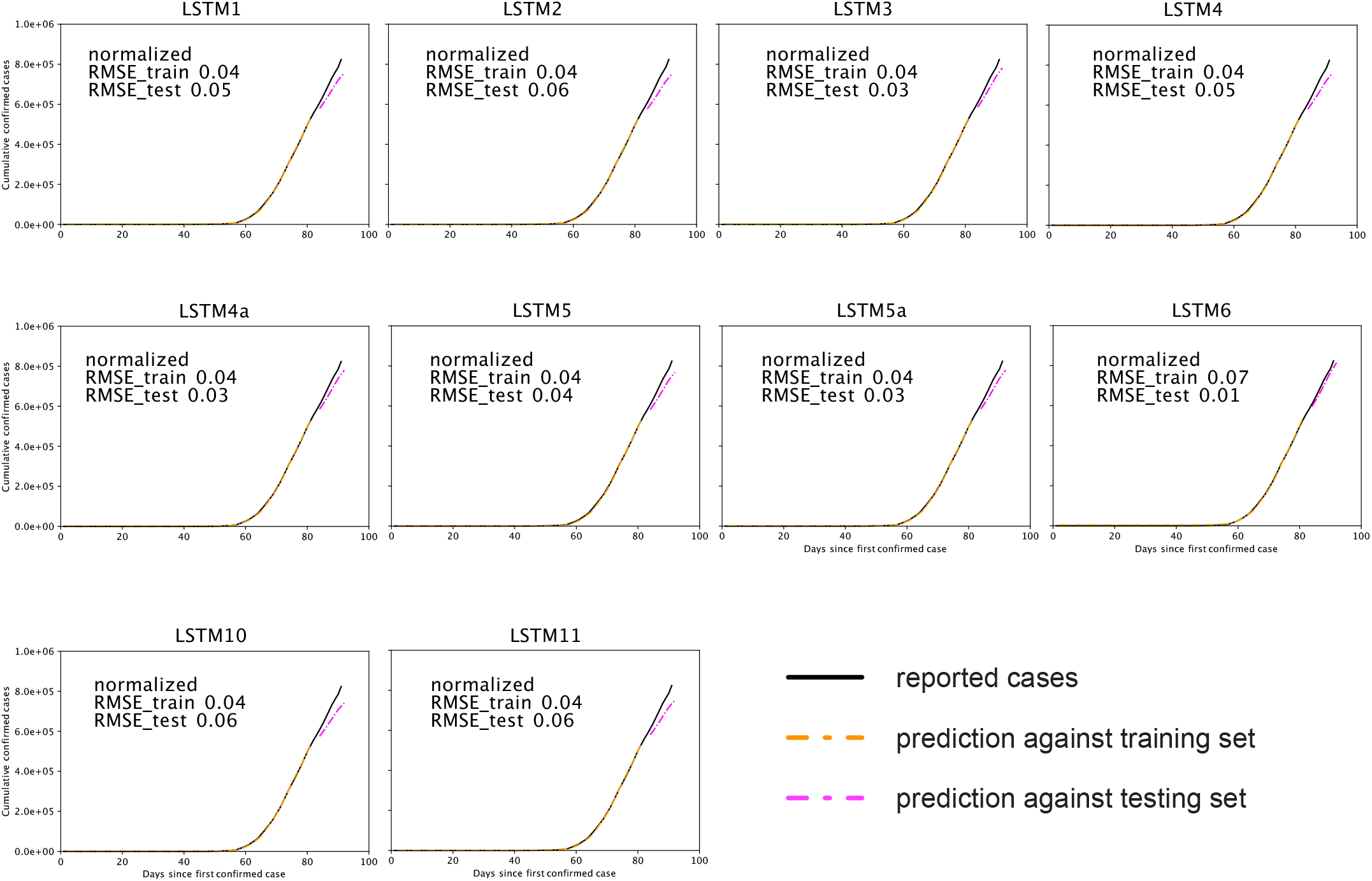
Prediction of cumulative patients in USA with slim LSTM RNN(s) using 82-day data.

## 5. Summary

This paper assesses full-range predictions of COVID-19 pandemic in USA with curve fitting using data of different lengths and compares short-term (8 to 10 days) predictions with curve fitting and 11 different LSTM recurrent neural networks. The full-range predictions using the latest data (up to April 28) suggests the pandemic in USA would end around the 160th day (June 30, 2020) since the first reported case and there would be over 1.3 × 10^6^ infections and 7.7 × 10^4^ deaths. Nevertheless, as the pandemic is evolving quickly on the daily basis and there are many variables (policies, people’s compliance to stay-home order etc.) that affect the real situation, one-time prediction of the ultimate date and scale might not be reliable enough. Therefore, it is advisable to fine tune the full-range predictions with the evolving data for the curve fitting method. In terms of short-term predictions, LSTM6 does the prediction with the highest accuracy among the standard LSTM RNN and 10 types of slim LSTM RNNs tested in this study. However, LSTM RNNs do not show advantage over curve fitting for this type of predictions. Curve fitting might be better to fit the true distribution of how COVID-19 infections “behave” because it does not overfit on the training set compared to RNN.

## Data Availability

Data source is located at Github repository https://github.com/CSSEGISandData/COVID-19, which is originally from Johns Hopkins University Coronavirus Resource.

## 6. Acknowledgement

We greatly appreciate the inspiration and help from professor Fathi S. Salem at Michigan State University. The computational calculation was supported by Google Compute Engine. Jupyter notebook for this study is hosted at https://github.com/zhuowenzhao/COVID19-prediction

## 7. Appendix

Gate equations parameter-reductions[1]:

LSTM1

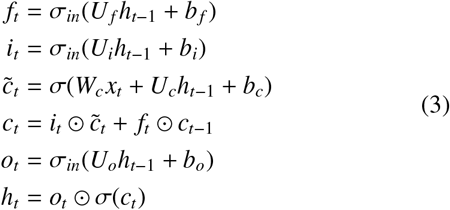

LSTM2

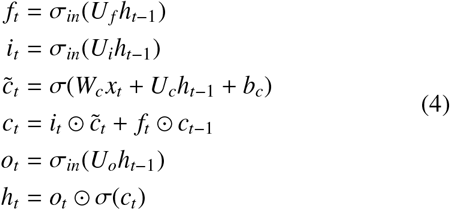

LSTM3

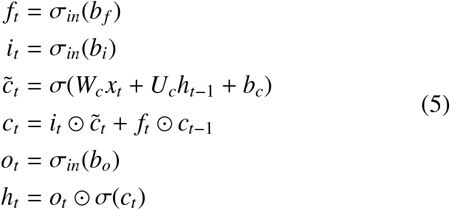

LSTM4

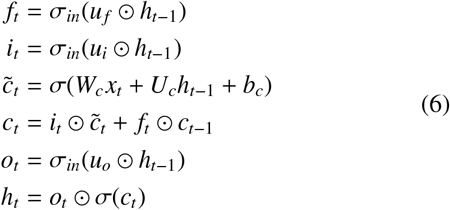

LSTM4a

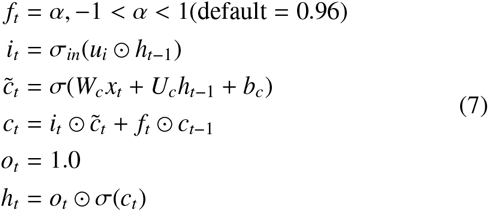

LSTM5

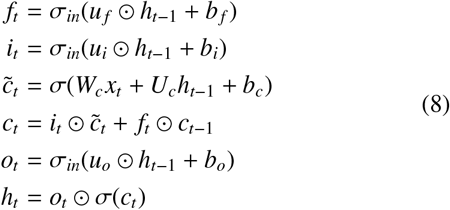

LSTM5a

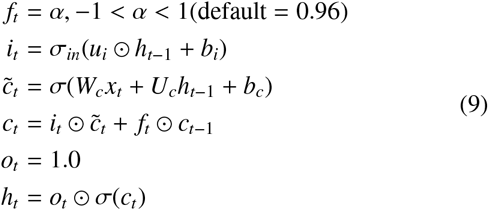

LSTM6

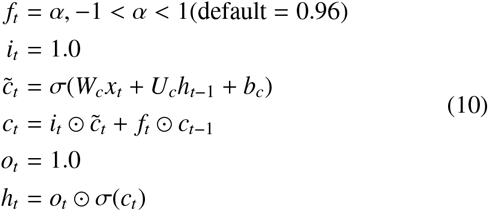

Memory cell equations parameter reductions:

LSTM10

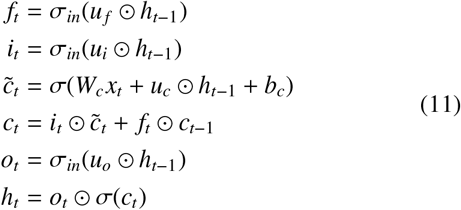

LSTM12

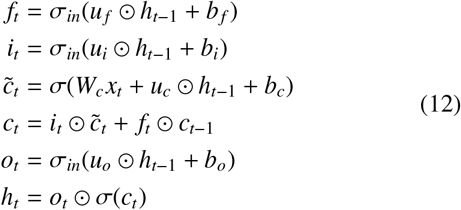

